# Determinants of mobility decline in nephrology-referred patients with CKD: a longitudinal cohort study

**DOI:** 10.1101/2022.03.30.22273207

**Authors:** Se Ri Bae, David A. Goodson, Chenoa R. Vargas, Tae Youn Kim, Gwenaelle Begue, Cynthia Delgado, Cassianne Robinson-Cohen, Jorge Gamboa, Jonathan Himmelfarb, Ian H. de Boer, Bryan Kestenbaum, Baback Roshanravan

## Abstract

**Background and Objective:** Chronic kidney disease (CKD) is associated with loss of muscle quality leading to mobility limitation and decreased independence. Identifying predictors of gait speed decline may help target rehabilitative therapies to those at highest risk of mobility impairment.

**Design, setting and participants, and measurements:** The current prospective cohort study recruited ambulatory patients with stage 1-4 CKD (eGFR 15-89 ml/min/1.73m^2^) from nephrology clinics. Predictors included demographic and clinical variables including GFR estimated using serum cystatin C. Outcomes were average change in gait speed (m/s) per year and inclusion in the top tertile of gait speed decline over 3 years. Linear mixed models and relative risk regression were used to estimate associations with annual gait speed changes and fastest tertile of decline.

**Results:** Among 213 participants, 81% were male, 22% were black and 43% had diabetes. Mean age was 57±13 years, median follow-up 3.15 years, mean baseline eGFRcysc 47.9±21ml/min/1.73 m^2^, and median baseline gait speed 0.95m/s [IQR 0.81, 1.10]. Lower baseline eGFRcysc was associated with more rapid loss of gait speed (−0.029 m/s/year [95% CI -0.042, -0.015] per 30 ml/min/1.73 m^2^ lower eGFR; *p*<0.001). Diabetes was associated with -0.024m/s/year faster change (95% CI -0.042, -0.007; p=.007). Lower eGFRcysc was associated with a 49% greater risk of rapid gait speed decline (IRR 1.49; 95% CI 1.11, 2.00, *p*=.008) after adjustment.

Prevalent cardiovascular disease and African American race were associated with a 45% greater (IRR 1.45; 95% CI 1.04, 2.01, *p*=.03) and 58% greater rate of rapid gait speed decline (IRR 1.58; 95% CI 1.09, 2.29, *p*=.02), respectively.

**Conclusions:** Among ambulatory, disability-free patients with CKD, lower eGFRcysc and diabetes status were associated with faster gait speed decline. Lower eGFRcysc, cardiovascular disease, and African American race were associated with rapid gait speed decline.

## Introduction

Chronic kidney disease (CKD) is a major public health issue that affects approximately 15% of the US adult population(1). CKD is associated with reduced skeletal muscle quality and impaired physical mobility that can result in frailty, falls and loss of functional independence(2–4). Hypothesized mechanisms underlying skeletal muscle dysfunction in CKD include atrophy of type II muscle fibers(5), metabolic acidosis(6,7), increased sympathetic tone(8), inflammation(9), retention of metabolic wastes(10) that lead to oxidative stress, insulin resistance, mitochondrial dysfunction and abnormal proteostasis(11).

Gait speed is a valid and clinically important indicator of mobility and functional limitation. In populations without kidney disease, gait speed measurements have high inter-rater reliability(12) and are associated with future risks of falls, hospitalizations, disability, and death(13). The loss of gait speed over time may be a particularly sensitive harbinger of subclinical disease. Among well-functioning older adults, declines in gait speed over two-years were strongly associated greater risks of all-cause mortality(14).

Given the clinical relevance of gait speed and the complex biological mechanisms contributing to its decline in kidney disease, we investigated the determinants of longitudinal gait speed decline among ambulatory patients with CKD not treated with dialysis.

## Methods

### Study Population

The Seattle Kidney Study (SKS) is a clinic-based prospective cohort study of non–dialysis-dependent CKD based in Seattle, WA. The SKS was designed to evaluate long-term complications of CKD, including cardiovascular disease, decline in kidney function, and changes in physical functioning. The SKS began recruiting in 2004 and enrolled 693 participants from outpatient nephrology clinics at the Veterans Affairs Puget Sound Health Center, Harborview Medical Center, and the University of Washington Medical Center. Eligibility criteria included age greater than 18 years and any stage of CKD not requiring dialysis defined by an estimated glomerular filtration rate (eGFR) <90 mL/min/1.73 m^2^ or the presence of albuminuria (urine albumin-creatinine ratio 30 mg/g from a 12-hour urine collection). Exclusion criteria included current or previous dialysis treatment, kidney transplantation, dementia, institutionalization, participation in a clinical trial, non–English speaking, or inability to undergo the informed consent process. Institutional review boards at the University of Washington and Veterans Affairs Puget Sound Health Care System approved the SKS in adherence with the Declaration of Helsinki and all participants provided written informed consent.

Among 693 SKS participants, we included 410 participants who had available gait speed measurements at their initial visit and at least one annual follow-up visit. To focus on functional individuals at baseline, we excluded 40 participants who had one or more disability in their activities of daily living (ADL) at baseline (self-reported difficulty in any 15 tasks of daily life including feeding, dressing, and bathing oneself(15,16)), 9 who were wheelchair dependent, and 52 who had severe lower extremity physical impairment defined by a Short Physical Performance Battery (SPPB) test score <8 (corresponding to less than the lowest tertile of scores in persons with CKD)(17)(Figure 1). SPPB score <8 has been associated with substantially greater likelihood of mobility disability and imminent risk of hospitalization or death(18,19).

**Figure 1.**
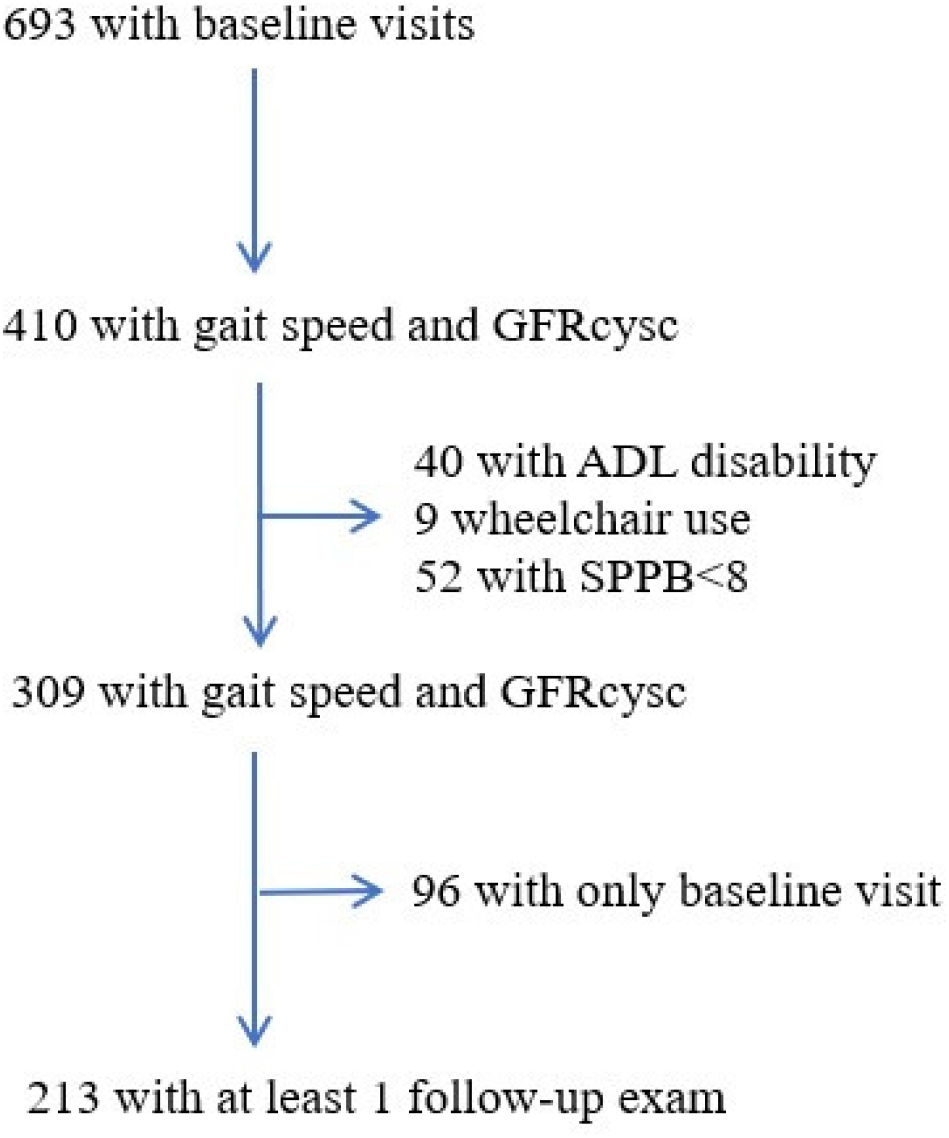
Participant flow diagram. Flow diagram of Seattle Kidney Study (SKS) participants included in the study. Abbreviations: eGFRcysc = Cystatin C based eGFR, ADL = activities of daily living, SPPB = Short Physical Performance Battery

### Gait Speed Assessment

The primary outcome was the absolute change in gait speed, measured in meters per second (m/s) per year. The secondary outcome was fast gait speed decline, defined as the highest tertile of decline (<-0.058 m/s/yr). Physical performance testing was conducted in the SKS from September 2006 until November 2015. Study coordinators performed two trials of gait speed, which were measured by asking participants to walk at their usual pace over a marked 4-meter course. As standard practice(20–22), the faster of the two trials was entered for analysis as it is believed to represent a participant’s normal speed after adjustment to the experimental setting. Prior observational studies of community dwelling older adults have defined meaningful change estimates for gait speed to be between 0.04 to 0.06 m/s(23).

### Covariate Measurements

We estimated GFR using the 2012 CKD-EPI cystatin C equation, which incorporates serum cystatin C concentrations, age, and sex (eGFRcysc)(24). Consistent with our previous studies, we selected cystatin C–based estimates of GFR due to the potential confounding influence of muscle mass on serum creatinine concentration(24,25). We also performed sensitivity analyses of eGFR using the combined creatinine-cystatin C based CKD-EPI equation (eGFRcr-cysc) for the benefit of practitioners who continue to rely on creatinine-based estimates. Serum samples were centrifuged for 20 minutes at 3,300 RPM, transferred to cryovials, and stored at -80°C. Laboratory personnel measured general chemistries using a Beckman-Coulter DXC autoanalyzer and serum cystatin C and C-reactive protein (CRP) concentrations using a Siemens Nephelometer (www.medical.siemens.com) that used a particle-enhanced immunonephelometric assay (N Latex Cystatin C)(25).

Calibration of cystatin C was performed using manufacturer’s standards along with daily quality controls. All samples were analyzed using assays from the same lot. Intra- and interassay coefficients of variation for cystatin C measured from 20 serum samples run in triplicate were 2.59% and 1.08%, respectively. We measured urinary albumin concentration by immunoturbimetry and urinary creatinine concentration by the modified Jaffé method.

Prevalent diseases were defined based on participant responses to SKS questionnaires, medication use, laboratory findings, and hospitalizations that occurred after initial SKS enrollment(16). Medication use was assessed by the inventory method at the Harborview and University of Washington sites and the electronic pharmacy database at the Veterans Affairs Puget site; missing medication data were completed by chart review. At each visit, study coordinators measured blood pressure and collected serum, plasma, and 12-hour timed urine samples on the same day as the gait speed assessment.

### Follow-up

Study coordinators contacted participants by telephone every 6 months and in-person through annual study examinations. Coordinators assessed vital status using medical record review, contact with family members, and the Social Security Death Index. Among the 213 participants, 44 participants had one year of follow-up, 40 had two years of follow-up, 40 had 3 years of follow-up, and 89 participants had 4 or more years of follow-up (Supplementary Table 1).

### Statistical Analysis

We described baseline characteristics comparing participants in each tertile of gait speed categorized into fastest, middle, and slowest designations over the entire duration of follow-up. Linear mixed models were constructed to estimate associations of demographic, clinical and laboratory characteristics with annual changes in gait speed. CKD severity was categorized in eGFRcysc 60 or greater, 40-59, 30-39, and <30 ml/min per 1.73m^2^. Models were adjusted for age, sex, self-reported race (binary variable, African American or not), height, weight, education, smoking, baseline gait speed, prevalent diabetes mellitus and prevalent cardiovascular disease. Given their biologically plausible confounding on the association of eGFR with change in gait speed, we included both fixed effects and interaction terms with time for age, sex, body weight, height and race in our model. A sensitivity analysis was performed adjusting for bicarbonate, hemoglobin, and phosphate. Association of gait speed change with eGFRcysc as a continuous variable in participants with eGFR<60 ml/min was also performed for sensitivity analysis.

We calculated the annual slope of decline for each person over the median follow up period of 3 years. Percent annual decline was determined from the slope of change from log transformed gait speed values and then determining subject specific mean percent difference in gait speed using the following formula ((e^β^)*100)-100. Relative risk regression was performed using multivariable generalized linear models with log link and gaussian distribution to calculate incidence rate ratios (IRR) for associations of demographic, clinical variables, and lab measures with the fastest tertile of gait speed decline. Demographic and clinical variables included GFRcysc, age, sex, height, weight, baseline gait speed, race (African American), education, current smoking status, diabetes, cardiovascular disease, and log transformed C-reactive protein (CRP). Multiple imputation using chained equations were performed for missing variables. Statistical analysis was performed using STATA version 17.

## Results

### Participant characteristics

Of the 213 SKS participants, 81% were male, 22% self-reported Black race, and 43% had a history of diabetes. The mean age was 57 ±13 years and the median follow-up was 3.2 ±1.8 years. The mean eGFRcysc was 48 ±21 ml/min/1.73 m^2^ and the median gait speed was 0.95 [IQR 0.81, 1.10] m/s at baseline. Annual gait speed changes among participants were normally distributed (Figure 2). Baseline characteristics including demographics, laboratory and clinical values, comorbidities, medications, and physical performance assessment are outlined in Table 1. Participants with the fastest gait speed decline were more likely be male and have diabetes and lower baseline eGFRcysc.

**Figure 2.**
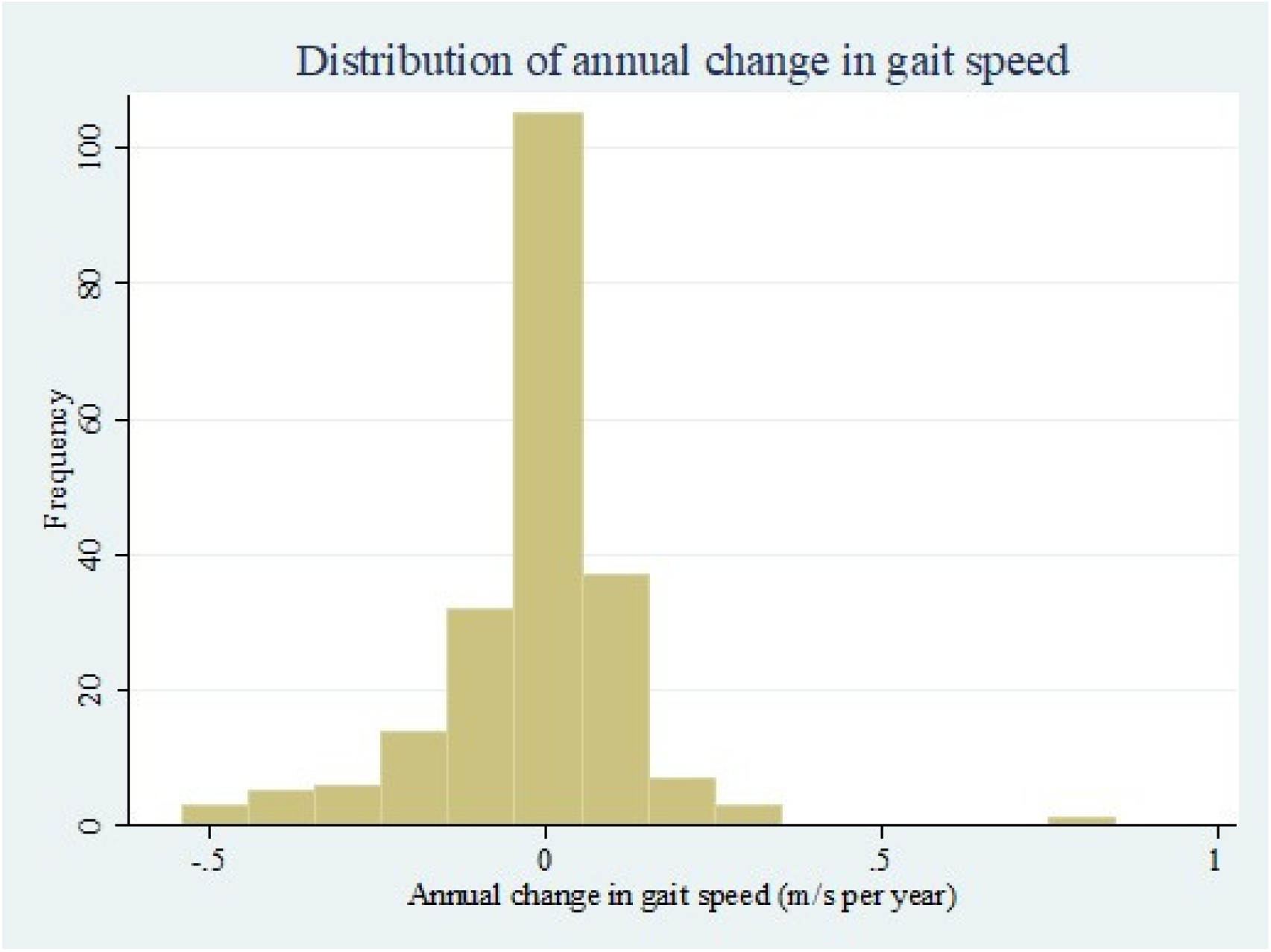
Distribution of annual change in gait speed. Annual changes in gait speed among participants revealed a normal distribution.

**Table 1.**
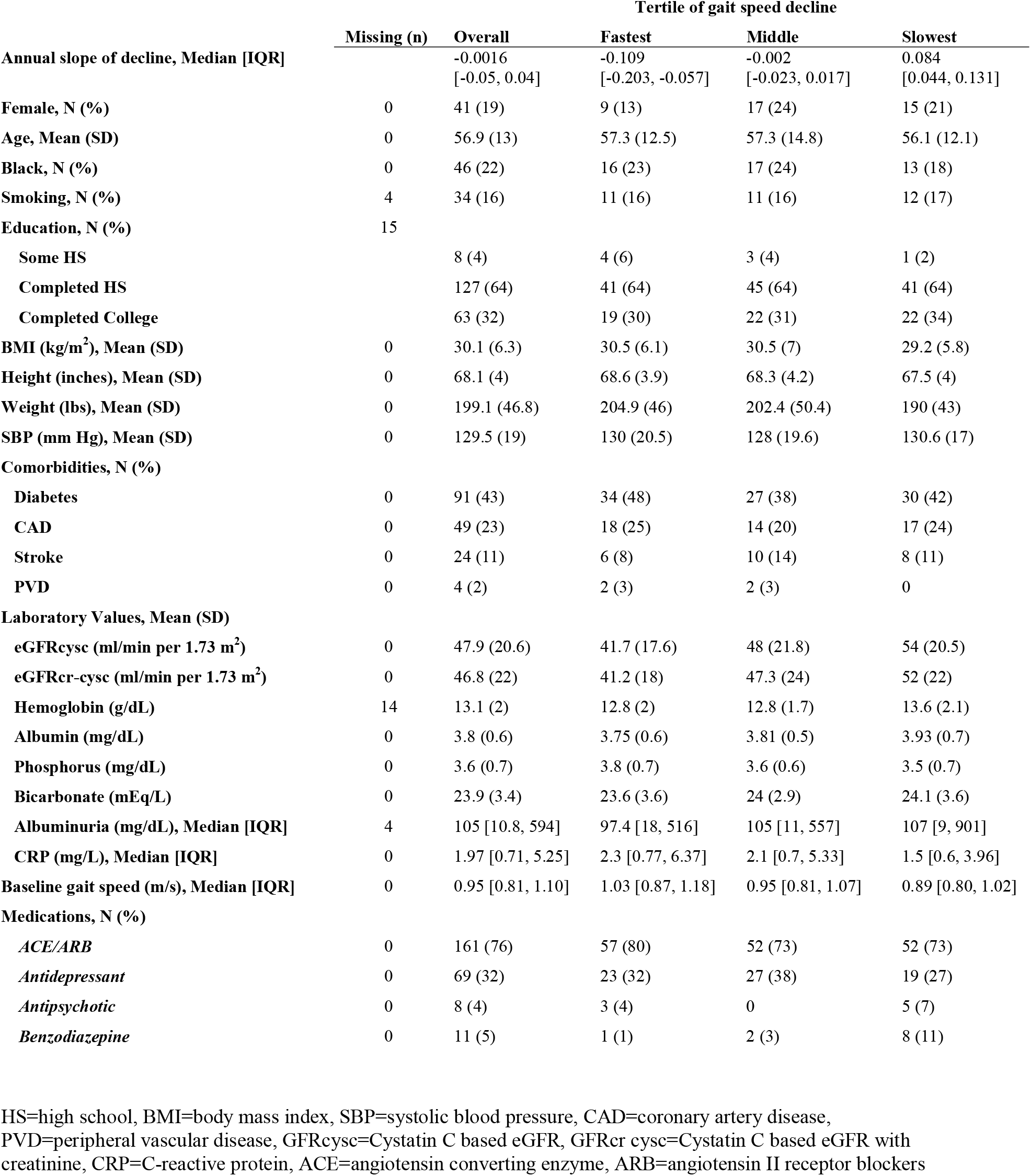
Baseline characteristics of study participants.

### Association of gait speed change with demographic and clinical variables

Lower baseline eGFRcysc was associated with faster declines in gait speed after adjustment for age, sex, race, height, weight, education, and smoking, and baseline gait speed (Table 2). Each 30 ml/min per 1.73m^2^ lower baseline eGFRcysc was associated with an estimated 0.029 m/s/year greater annual decline in gait speed (95% CI -0.042, -0.015; *p*<0.001) after adjustment. Gait speed declined more rapidly in lower eGFRcysc categories (Table 2).

**Table 2.** Associations of clinical characteristics with change in gait speeds.

|  | Mean (SD)<br>baseline gait<br>speed (m/s) | Mean (SD)<br>annual change<br>(m/s per year) | Model 1<br>Adjusted annual<br>change (95% CI) | Model 2<br>Adjusted annual<br>change (95% CI) | Sensitivity Analysis<br>Adjusted annual change<br>(95% CI) |
| --- | --- | --- | --- | --- | --- |
| **eGFRcysc** |  |  |  |  |  |
| ≥60 (n=52) | 1.00 (0.19) | 0.022 (0.119) | Reference | Reference | Reference |
| 40-59 (n=76) | 0.96 (0.23) | 0.002 (0.143) | -0.028 (-0.049, -0.007) | -0.034 (-0.054, -0.013) | -0.022 (-0.045, 0.003) |
| 30-39 (n=46) | 0.92 (0.17) | -0.049 (0.151) | -0.034 (-0.062, -0.005) | -0.040 (-0.067, -0.013) | -0.033 (-0.062, -0.003) |
| <30 (n=39) | 0.98 (0.21) | -0.069 (0.152) | -0.077 (-0.108, -0.046) | -0.078 (-0.107, -0.049) | -0.049 (-0.082, -0.016) |
| **Per 30 ml/min lower**<br>**eGFRcysc** |  |  | -0.027 (-0.041, -0.012) | -0.029 (-0.042, -0.015) | -0.024 (-0.039, -0.009) |
| **Age (per 1 year)** |  |  | 0.001 (-0.000, 0.002) | 0.001 (0.000, 0.002) | 0.001 (0.000, 0.002) |
| **Sex (female)** |  |  | 0.001 (-0.035, 0.036) | -0.001 (-0.034, 0.033) | 0.004 (-0.030, 0.037) |
| **Height (per 1 inch)** |  |  | -0.001 (-0.004, 0.003) | -0.001 (-0.004, 0.002) | -0.002 (-0.005, 0.002) |
| **Weight (per 10 lbs)** |  |  | -0.002 (-0.003, 0.000) | -0.001 (-0.003, 0.000) | -0.001 (-0.003, 0.000) |
| **African American race** |  |  | -0.013 (-0.031, 0.005) | -0.018 (-0.035, -0.000) | -0.014 (-0.030, 0.002) |
| **No Diabetes (n=122)** | 0.97 (0.21) | -0.008 (0.154) |  | Reference | Reference |
| **Diabetes (n=91)** | 0.96 (0.20) | -0.029 (0.132) |  | -0.024 (-0.042, -0.007) | -0.024 (-0.042, -0.007) |
| **Hemoglobin (per 1gm/dL)** |  |  |  |  | 0.006 (0.000, 0.011) |
| **P for continuous eGFRcysc** |  |  | 0.001 | <0.001 | 0.002 |
| **P value for age** |  |  | 0.093 | 0.020 | 0.021 |
| **P value for sex** |  |  | 0.964 | 0.976 | 0.836 |
| **P value for height** |  |  | 0.758 | 0.579 | 0.333 |
| **P value for weight** |  |  | 0.053 | 0.076 | 0.094 |
| **P value for African**<br>**American race** |  |  | 0.143 | 0.044 | 0.097 |
| **P value diabetes** |  |  |  | 0.007 | 0.007 |
| **P value for hemoglobin** |  |  |  |  | 0.034 |

Model 1: Age, sex, race (African American vs. other), height, weight, education, smoking, baseline gait speed
Model 2: + Diabetes, Any CVD (CAD, PVD, stroke)
Sensitivity Analysis: + Bicarbonate, hemoglobin, phosphate. Note hemoglobin specified as the main effect and interaction term with time in model eGFRcysc=Cystatin C based eGFR, CVD=cardiovascular disease, CAD=coronary artery disease, PVD=peripheral vascular disease

Diabetes was independently associated with a -0.024 m/s/year (95% CI -0.042, -0.007; *p* =.007) faster decline in gait speed compared to no diabetes. Findings were consistent for kidney function measured using the combined eGFRcr-cysc equation (Supplemental table 2).

Sensitivity analysis adjusting for hemoglobin attenuated the association of eGFRcysc with gait speed decline by 17% (Table 2). There was no association of serum phosphorus or bicarbonate with gait speed decline. Additionally, when restricting analysis to participants with eGFRcysc<60 (n=149), each 30 ml/min per 1.73m^2^ lower eGFRcysc was associated with an estimated mean decline of 0.032 m/s/year (95% CI -0.061, -0.003; *p*=0.03) after adjustment for height, weight, race, education, smoking, baseline gait speed, and comorbidities. The association for eGFRcr-cysc<60 (n=147) was -0.038 m/s/year (95% CI -0.067, -0.009; *p*=0.01) after adjustment.

### Associations with fastest tertile of gait speed decline over the median follow-up period

The association of eGFRcysc with gait speed decline was confirmed in our secondary analysis examining the associations of demographic and clinical variables with rapid gait speed decline over the median follow-up period of 3.15 years [IQR 1.97, 4.3] (Table 3). Rapid gait speed decline was defined as the fastest tertile of gait speed decline (<-0.058 m/s/yr; median 17.5% per year decrease in gait speed [IQR 12, 28]). Each 30 ml/min per 1.73m^2^ decrement in eGFRcysc was associated with a 49% greater incidence of rapid gait speed decline (IRR 1.49; 95% CI 1.11, 2.00, *p*=.008) after adjustment. Prevalent cardiovascular disease was associated with a 45% greater incidence of rapid gait speed decline (IRR 1.45; 95% CI 1.04, 2.01, *p*=.03). African American race was associated with a 58% greater incidence of rapid gait speed decline compared to non-African Americans (IRR 1.58; 95% CI 1.09, 2.29, *p*=.02) (Table 3). In an adjusted model using the eGFRcr-cysc estimation equation, lower eGFR (IRR 1.44; 95% CI 1.07, 1.93, p=0.02), presence of cardiovascular disease (IRR 1.46, 95% CI 1.05, 2.04; p=0.03) and African American race (IRR 1.50; 95% CI 1.03, 2.19; p=0.03) remained significant risk factors associated with risk of rapid gait speed decline (Supplementary Table 3). In sensitivity analyses including only participants with eGFRcysc<60, each 30 ml/min per 1.73m^2^ lower eGFRcysc was associated with a 1.69 fold higher IRR of rapid decline (95% CI 1.20, 2.44; p<0.01). In this subset, 57 demonstrated rapid decline (36%) and 102 (64%) did not.

**Table 3.** Associations of clinical characteristics with rapid gait speed decline over 3 years of follow-up.

|  | Model 1 |  | Model 2 |  |
| --- | --- | --- | --- | --- |
|  | IRR (95% CI) | P-value | IRR (95% CI) | P-value |
| **eGFR<sub>cysc</sub> (per 30 ml/min per 1.73m<sup>2</sup> decrement)** | 1.46 (1.10, 1.95) | 0.009** | 1.49 (1.11, 2.00) | 0.008** |
| **Age (per 1 yr older)** | 1.01 (0.99, 1.03) | 0.332 | 1.00 (0.99, 1.02) | 0.634 |
| **Height (inches)** | 1.03 (0.95, 1.11) | 0.477 | 1.03 (0.95, 1.13) | 0.466 |
| **Weight (lbs)** | 1.00 (1.00, 1.01) | 0.332 | 1.00 (1.00, 1.01) | 0.229 |
| **Female gender** | 0.75 (0.33, 1.73) | 0.503 | 0.80 (0.31, 2.08) | 0.651 |
| **African American** | 1.46 (1.04, 2.04) | 0.030* | 1.58 (1.09, 2.29) | 0.017* |
| **Education**<br>**(Greater than HS vs HS or less)** | 0.86 (0.53, 1.39) | 0.539 | 0.89 (0.60, 1.33) | 0.577 |
| **Current smoking** | 1.78 (1.00, 3.16) | 0.049 | 1.60 (0.93, 2.75) | 0.087 |
| **Diabetes** |  |  | 1.15 (0.81, 1.63) | 0.440 |
| **Cardiovascular disease** |  |  | 1.45 (1.04, 2.01) | 0.027* |
| **CRP (per 1 log mL/L greater)** |  |  | 1.04 (0.91, 1.19) | 0.590 |

Rapid gait speed decline defined as the fastest tertile (<-0.058 m/s/yr; median 17.5% per year decrease in gait speed [IQR 12, 28])
GFR<sub>cysc</sub>=Cystatin C based eGFR, HS=high school, CRP=C-reactive protein; \*\* $p<0.01$ , \* $p<0.05$

## Discussion

Among ambulatory and disability-free patients referred to nephrology clinics with stage 1-4 CKD, lower baseline eGFRcysc and diabetes were associated with faster decline in gait speed over time after adjusting for demographics characteristics and comorbidities. Diabetes and baseline hemoglobin were independently associated with faster decline in gait speed. In addition, lower baseline eGFRcysc, self-reported African American race, and prevalent cardiovascular disease were associated with clinically meaningful rapid gait speed decline. Associations between kidney function by eGFR with gait speed decline appeared greater when estimates of GFR were not adjusted for race.

For the first time, we demonstrate that diabetes and lower eGFR are associated with faster declines in gait speed over time in patients with CKD without severe physical impairment or ADL disability. Our study adds to prior cross-sectional studies in populations with CKD indicating diabetes as a determinant of slower gait speed (26,27). We also found that worsening kidney disease, measured as eGFRcysc, is independently associated with faster gait speed decline. Lower kidney function has been shown to adversely impact muscle metabolic health and mobility in patients with CKD(7) and in patients living with diabetes(28). Specifically, among patients with diabetes, lower kidney function is associated with more pronounced muscle mitochondrial dysfunction, contributing to more severe declines in performance of daily physical activities including walking, climbing stairs, or picking up objects from the ground. The current study builds upon a growing base of evidence implicating mitochondrial dysfunction in CKD-associated muscle impairment(9,29).

Low hemoglobin attenuated the association of eGFRcysc and diabetes with gait speed decline, suggesting the potential impact of anemia on physical function in CKD. Anemia is believed to be a biomarker of lean body mass loss, inflammation, and skeletal muscle abnormalities leading to impaired physical function(30). Previous studies have reported that CKD and anemia are independently associated with reduced physical performance(31,32); however, the relationship between anemia, CKD, and physical function is complex due to overlapping cardiovascular comorbidities and implications of aging. Therefore, the effectiveness of treating anemia in CKD to improve physical function remains inconclusive.

Self-reported African American race, lower baseline eGFRcysc, and cardiovascular disease were significantly associated with increased risk of rapid gait speed decline. Prior studies identified self-reported African American race as a risk marker for frailty. In the Cardiovascular Health Study, Hirsch et al. (2006) found a fourfold greater odd of frailty in African Americans compared to whites(33). Among African American older adults, patients with diabetes were more likely to be frail and have worse SPPB performance and increased risk of mortality compared to those without diabetes(34), further reinforcing the role of diabetes in declining physical performance in this high-risk population. These disparities warrant further dissection of the impact of social determinants and allosteric load on increased risk of frailty and mobility impairment(35).

Gait speed has been considered a vital test capturing deficits in neuromuscular(36,37) and cognitive function (36–38) in addition to skeletal muscle health across multiple populations. A systematic review by Shen et al. (2017) found that decreased kidney function was strongly associated with cognitive decline and that vascular disease is likely an important mediator(41). Gait abnormalities in CKD may be associated with cognitive-motor interference, with lower eGFR having shown to be associated with greater gait abnormalities during walking-while-talking tasks among community-dwelling, nondisabled adults with CKD(41). The potential independent contribution of cognitive impairment on gait speed in CKD is not captured in our study and remains yet to be elucidated. Nonetheless, utilizing gait speed as an assessment of mobility can facilitate early intervention with physical rehabilitative therapies to reverse physical impairment in CKD.

Our study supports previous findings that demonstrate a strong connection between renal disease and physical impairment(42). Previous studies have found that the presence of CKD is associated with lower gait speeds and that physical function is further reduced in more advanced CKD(42,43). In our study, participants with the fastest tertile of gait speed decline had a median gait speed change of -0.109 m/s/year, which is comparable to the minimal clinically important difference identified over a broad range of diseases(44,45). In a longitudinal cohort study of ambulatory older adults with CKD, each 0.1 m/s decrement in gait speed at baseline was associated with a 26% higher risk for all-cause mortality(46). Our study reveals meaningful change in gait speed among ADL disability-free patients with CKD with implications for poor clinical outcomes.

Limitations of our study include the low prevalence of female participants in the cohort, which precludes estimates of sex-specific changes in gait speed over time. Potential differences in survivorship as a form of non-random selection for the biometric data included in the study may bias our results. However, we expect this would bias results toward more conservative estimates of association. Unmeasured characteristics among those with lower eGFR may contribute to residual confounding in the association of eGFRcysc with gait speed decline.

Strengths of our study include our relatively large size of 213 nephrology-referred ambulatory CKD patients without overt mobility disability, longitudinal measures of gait speed, and use of more accurate estimation of GFR by cystatin C. Estimates of glomerular filtration rates were based on serum cystatin C concentrations and not serum creatinine concentrations, which may be influenced by non-GFR determinants (i.e. diet, medications, and muscle mass). The cystatin C-based equation also does not include a race coefficient(24), further bolstering our findings that African Americans are at a higher risk of rapid gait speed decline even after adjusting for education, an indirect measure of socioeconomic status. eGFRcysc also better captured gait speed decline, as evidenced by the relative attenuated estimates of association using eGFRcr-cysc (Supplementary Table 2). In comparison to a prior study demonstrating an association between CKD and mobility decline in community-dwelling older adults enrolled in the Framingham study(48), our study included more frequent gait speed assessments in a cohort of participants with a broader age range who were free of both severe lower extremity impairment and ADL disability.

In summary, we demonstrated that among ambulatory, ADL disability-free patients with CKD, lower kidney function by eGFRcysc, diabetes, and anemia were associated with faster gait speed declines. Furthermore, the risk markers including African American race, lower baseline eGFRcysc and cardiovascular disease were associated with higher risk for rapid decline in gait speed. These associations can inform clinical management of CKD by facilitating mobility prognostication and by highlighting the role of glycemic control and cardiovascular disease management in disease progression. Larger studies are necessary to examine cut-points for gait speed decline associated with mortality, and further research is needed to understand the role of African American race, anemia, and cognitive decline in mobility disability in CKD.

## Data Availability

All data produced in the present study are available upon reasonable request to the authors

