## Supplementary Material for "Determinants of mobility decline in nephrology-referred patients with CKD: a longitudinal cohort study"

### Supplementary Materials

**Supplementary Table 1. Participant demographics by number of follow-up years**

| | | Maximum years of follow-up | | | | | |
|---|---|---|---|---|---|---|---|
| | **Missing** | **Baseline only** | **1 year** | **2 years** | **3 years** | **4 or more years** | **Study Cohort** |
| | | n=96 | n=44 | n=40 | n=40 | n=89 | n=213 |
| Female, no (%) | 0 | 25 (26) | 5 (11) | 9(23) | 8 (20) | 19 (21) | 41 (19) |
| Age (yrs), mean (SD) | 0 | 56.4 (13) | 59.9 (12) | 53.4 (14) | 58.8 (12) | 56.2 (13) | 56.9 (13) |
| Black, no (%) | 0 | 28 (29) | 12 (27) | 8 (20) | 6 (15) | 20 (22) | 46 (22) |
| Smoking, no (%) | 5 | 26 (27) | 10 (23) | 9 (23) | 4 (10) | 11 (13) | 34 (16) |
| Education, no (%) | 25 | | | | | | |
| Some Highschool | | 7 (8) | 3 (7) | 1 (3) | 1 (3) | 3 (4) | 8 (4) |
| Completed Highschool | | 69 (69) | 27 (68) | 21 (60) | 26 (68) | 53 (62) | 127 (64) |
| Completed College | | 20 (23) | 10 (27) | 13 (37) | 11 (29) | 29 (34) | 63 (32) |
| BMI, mean (SD) | 0 | 31.1 (8) | 28.5 (6.4) | 29.5 (5.7) | 31.8 (7.3) | 30.3 (6) | 30.1 (6.3) |
| SBP, mmHg mean (SD) | 0 | 138 (24) | 132 (18) | 129 (22) | 130 (17) | 128 (19) | 130 (19) |
| DM, no (%) | 0 | 48 (50) | 18 (37) | 19 (48) | 17 (43) | 37 (45) | 91 (43) |
| CAD, no (%) | 0 | 37 (38) | 12 (24) | 10 (25) | 10 (25) | 17 (20) | 49 (23) |
| eGFRcysc, mean(SD) | 0 | 40.1 (23) | 41.9 (18) | 42.3 (21) | 45.7 (18) | 54.4 (21) | 47.9 (21) |
| eGFRcr-cysc, mean (SD) | 0 | 40.2 (26) | 40.8 (18) | 40.9 (22) | 45.4 (19) | 52.2 (23) | 46.8 (22) |
| Hemoglobin (gm/dL), mean (SD) | 20 | 12.6 (1.9) | 13 (2) | 12.8 (2) | 12.9 (2) | 13.3 (2) | 13.1 (2) |
| Phosphorus (mg/dL), mean (SD) | 0 | 4 (0.9) | 3.8 (0.7) | 3.9 (0.7) | 3.7 (0.6) | 3.4 (0.6) | 3.6 (0.7) |
| Albuminuria (mg/dL) median (IQR) | 5 | 217 (23, 1265) | 106 (22, 929) | 324 (68, 1011) | 114 (13, 696) | 45 (15, 689) | 105 (10.8, 594.3) |
| Frail, no (%) | 57 | 11 (13) | 6 (17) | 3 (10) | 2 (6) | 3 (4) | 14 (8) |
| Gait speed (m/s), mean (SD) | 0 | 0.94 (0.21) | 0.93 (0.20) | 0.97 (0.21) | 0.97 (0.25) | 0.98 (0.18) | 0.97 (0.2) |

HS=high school, BMI=body mass index, SBP=systolic blood pressure, DM=diabetes mellitus, CAD=coronary artery disease, GFRcysc=Cystatin C based eGFR, GFRcr-cysc=Cystatin C based eGFR with creatinine, PTH=parathyroid hormone. Frailty was defined using slight modifications of criteria originally established by Fried et al.[1,2]

**Supplementary Table 2. Associations with annual changes in gait speed using eGFR with combined cystatin and creatinine**

| eGFRcr-cysc | Mean (SD) | Model 1 Adjusted annual change (95% CI) | Model 2 Adjusted annual change (95% CI) | Sensitivity Analysis Adjusted annual change (95% CI) |
|---|---|---|---|---|
| ***≥60 (n=52)*** | 1.02 (0.22) | Reference | Reference | Reference |
| ***40-59 (n=75)*** | 0.96 (0.20) | -0.016 (-0.039, 0.008) | -0.016 (-0.039, 0.007) | -0.012 (-0.033, 0.009) |
| ***30-39 (n=38)*** | 0.89 (0.17) | -0.023 (-0.051, 0.005) | -0.032 (-0.063, -0.002) | -0.016 (-0.054, -0.022) |
| ***<30 (n=48)*** | 0.98 (0.20) | -0.044 (-0.077, -0.011) | -0.044 (-0.074, -0.014) | -0.034 (-0.065, -0.003) |
| **Per 30 ml/min per 1.73m$^2$ eGFRcr-cysc** | | -0.016 (-0.031, -0.002) | -0.019 (-0.032, -0.006) | -0.016 (-0.029, -0.003) |
| **Age (per 1 year)** | | 0.001 (-0.000, 0.002) | 0.001 (-0.000, 0.002) | 0.001 (-0.000, 0.002) |
| **Sex (female)** | | 0.004 (-0.033, 0.042) | 0.003 (-0.032, 0.039) | 0.009 (-0.025, 0.044) |
| **Height (per 1 inch)** | | -0.000 (-0.004, -0.003) | -0.001 (-0.004, 0.003) | -0.002 (-0.005, 0.002) |
| **Weight (per 10 lbs)** | | -0.002 (-0.004, 0.000) | -0.001 (-0.003, 0.000) | -0.002 (-0.003, 0.000) |
| **African American race** | | -0.011 (-0.029, 0.008) | -0.015 (-0.033, 0.003) | -0.012 (-0.028, 0.004) |
| **Diabetes** | | | -0.024 (-0.043, -0.005) | -0.023 (-0.041, -0.005) |
| **Hemoglobin (per 1gm/dL)** | | | | 0.008 (0.003, 0.013) |
| **P for continuous GFRcysc** | | 0.026 | 0.004 | 0.016 |
| **P value for age** | | 0.219 | 0.075 | 0.051 |
| **P value for sex** | | 0.824 | 0.866 | 0.597 |
| **P value for height** | | 0.859 | 0.684 | 0.354 |
| **P value for weight** | | 0.081 | 0.101 | 0.088 |
| **P value for African American race** | | 0.263 | 0.109 | 0.135 |
| **P value diabetes** | | | 0.012 | 0.012 |
| **P value for hemoglobin** | | | | 0.002 |

Model 1: Age, sex, race (African American vs. other), height, weight, education, smoking, baseline gait speed
Model 2: + DM, Any CVD (CAD, PVD, stroke)
(Sensitivity Analysis): + Bicarbonate, hemoglobin, phosphate. Note hemoglobin is the main effect and interaction term with time
GFRcr-cysc=Cystatin C based eGFR with creatinine, DM=diabetes mellites, CVD=cardiovascular disease, CAD=coronary artery disease, PVD=peripheral vascular disease

**Supplementary Table 3. Associations with fastest tertile of decline in gait speed over 3 years of follow-up using eGFR with combined cystatin and creatinine**

| | **Model 1** | | **Model 2** | |
|---|---|---|---|---|
| | **IRR (95% CI)** | **P-value** | **IRR (95% CI)** | **P-value** |
| **eGFRcr-cysc (per 30 ml/min per 1.73m$^2$ lower)** | 1.39 (1.06, 1.83) | 0.018* | 1.44 (1.07, 1.93) | 0.016* |
| **Age (per 1 yr older)** | 1.01 (0.99, 1.03) | 0.357 | 1.00 (0.98, 1.03) | 0.721 |
| **Height (in)** | 1.04 (0.96, 1.12) | 0.388 | 1.04 (0.95, 1.13) | 0.385 |
| **Weight (lbs)** | 1.00 (1.00, 1.01) | 0.417 | 1.00 (1.00, 1.01) | 0.302 |
| **Female gender** | 0.76 (0.33, 1.76) | 0.518 | 0.80 (0.31, 2.11) | 0.659 |
| **African American** | 1.37 (0.97, 1.94) | 0.077 | 1.50 (1.03, 2.19) | 0.034* |
| **Education (Greater than HS vs HS or less)** | 0.85 (0.52, 1.38) | 0.501 | 0.89 (0.59 1.32) | 0.552 |
| **Current smoking** | 1.81 (1.01, 3.25) | 0.047* | 1.61 (0.93, 2.78) | 0.088 |
| **Diabetes** | | | 1.16 (0.80, 1.69) | 0.427 |
| **Any cardiovascular disease** | | | 1.46 (1.05, 2.04) | 0.025* |
| **Log CRP (per 1 log mg/L)** | | | 1.04 (0.90, 1.19) | 0.595 |

Fastest tertile of gait speed decline defined as a median 17.5% per year decrease in gait speed [IQR 12, 28.3]
GFRcr-cysc=Cystatin C based eGFR with creatinine, HS=high school, CRP=C-reactive protein; ***p*<0.01, **p*<0.05
